# Revisiting Role of Bilateral Ligation of Internal Iliac Arteries and Preperitoneal Pelvic Packing for Hemorrhage Control in Patients with Pelvic Injuries in Resource Constraint Settings

**DOI:** 10.1101/2020.07.11.20151357

**Authors:** Dinesh Kumar Bagaria, Majid Anwer, Narendra Choudhary, Abhinav kumar, Pratyusha Priyadarshini, Niladri Banerjee, Junaid Alam, Amit Gupta, Biplab Mishra, Sushma Sagar, Subodh Kumar

## Abstract

**Background:** Since the description of bilateral ligation of internal iliac arteries (BLIIA) and preperitoneal pelvic packing (PPP) for haemorrhage control in pelvic injury patients, multiple reports have been published advocating its use with acceptable outcomes. We analyzed our experience with this technique in a setting where the facility of hybrid Operating room for simultaneous angioembolisation is not available.

**Material and Methods:** We prospectively analysed data of sixty-six patients who presented in a state of unresponsive shock with pelvic fracture between January 2014 and September 2019. After initial resuscitation, they all underwent BLIIA with PPP as part of damage control surgery.

**Results:** Out of 66 patients, 55 were male. The mean age was 36.12 years. All patients sustained blunt trauma, with road traffic injuries being the most common mechanism involving 65 % of the patients followed by fall from height. The mean systolic blood pressure at the time of surgery was 77 ±34.46mm Hg. Median packed red blood cell transfusion in the first 24 hours was 8.5 units with IQR of 6-12. The hemorrhage related mortality was 48%.

**Conclusion:** BLIIA with PPP may be considered as a viable treatment option in hemodynamically unstable patients with pelvic injuries in resource constraint facilities.

## Introduction

Hemorrhagic shock is the second most common cause of death in trauma patients after head injury (1). Standardized management protocols such as Advanced Trauma Life Support (ATLS) protocol and massive hemorrhage protocol (MHP) have resulted in a considerable improvement in the outcomes of trauma patients with hemorrhagic shock. However, still, approximately one-third of all hemorrhage related deaths following blunt trauma are preventable (2). Uncontrolled pelvic bleeding is a significant contributor to these preventable deaths (3). Overall mortality from pelvic injury is very high. It ranges from 40% to 90% among patients who present with features of hemorrhagic shock in the emergency department (ED) (4). The outcome of patients with pelvic injuries is not only determined by the anatomical and physiological parameters but also by the available resources and timely interventions. Immediate management of these patients consists of hemostatic resuscitation, pelvic circumferential compression device (PCCD) application in ED followed by measures for definitive control of bleeding. Angiographic embolization with or without preperitoneal pelvic packing (PPP) has been recommended by the World Trauma Association as the procedure of choice for definitive control of hemorrhage (5).

In low and middle-income countries (LMICs), there is a dearth of dedicated trauma care facilities and are responsible for 93 % of injury-related deaths worldwide (6). In these resource constraint settings, advanced resources such as an angiography suite for angioembolization, expertise, and their timely availability are hard to find. Such situations demand alternative viable and effective methods for definitive control of pelvic hemorrhage. Bilateral ligation of the internal iliac arteries (BLIIA) with PPP has been shown to be a rapid and effective alternative method of hemorrhage control in massive pelvic bleeding (7-8).

We reviewed our experience with BLIIA and PPP with a hypothesis that this technique is a viable option in resource constraint settings.

## Material and Methods

This study was performed at a standalone level 1 Trauma Center in India. Prospectively maintained data of patients who underwent BLIIA with PPP for hemorrhage control in case of isolated or associated severe pelvic trauma between January 2014 and September 2019 were reviewed.

All patients with pelvic trauma were managed as per ATLS protocol in ED, and a focused assessment by sonography for trauma (FAST) examination, and plain radiography of the chest and pelvis were done. A PCCD using a bed sheet was applied to all the patients having clinical or radiological features of pelvic fracture. Based on their response to initial resuscitation with fluids and blood products and findings on FAST, further course of management was decided. All non-responder patients with pelvic trauma were shifted to the operating room (OR) for damage control laparotomy. BLIIA and PPP were performed in case of significant bleeding from the pelvic fractures along with a thorough exploration of the peritoneal cavity. Various other surgical procedures were also combined in patients who had associated intra-abdominal injuries.

### Technique

After midline exploratory laparotomy, a quick intraperitoneal survey is being done to rule out any other significant source of bleeding. Once other sources ruled out, operating surgeon identifies the common iliac artery and its bifurcation by palpating arterial pulsation starting from aortic bifurcation over posterior peritoneum. On left side, mobilization of sigmoid colon might be needed. At the common iliac artery bifurcation posterior peritoneum is being opened taking care of overlying ureter and pushing it laterally. The proximal 3-4 cm of internal iliac artery is mobilized and is ligated at two sites in continuity without dividing. Same procedure is being repeated on other side. Once the BLIIA done, Space of Retzius is being approached by manually separating peritoneum from inner aspects of pubic symphysis and pelvic ring. Two to three surgical pads are packed on either side of bladder from posterior to anterior. After the procedure, abdomen is being closed or mesh laparostomy is being made depending upon situation.

The demographic data, mechanism of injury, hemodynamic status, associated injuries, injury severity scores, laboratory parameters, and operative details were reviewed. Details of blood transfusions were also recorded. All patients were followed up during their stay in the hospital, and their postoperative complications and final outcomes were noted.

Standard statistical analysis was done. Data are reported as means (SD) and medians (IQR). Student t-test for univariate variable and Chi-square test were used to compare the means. All statistical analyses were performed using SPSS (IBM, Version 23). A p-value< 0.05 was considered significant.

## Results

A total of 66 trauma patients were found eligible to be included in the study. Fifty-five patients (83.3 %) were males. The mean age was 36.12 years (SD, ± 12.97). All patients sustained blunt trauma, with road traffic injuries being the most common mechanism in 65 % of patients followed by fall from height. Majority of these patients (92.4%) presented with features of at least class II hemorrhagic shock (HS). The median injury severity score (ISS) was 30 (IQR: 26-36). The median length of stay (LOS) of these patients in the intensive care unit (ICU) was 2.5 (IQR: 1-11) days and in the hospital was 3.5 (IQR: 1-19) days. (Table −1)

Most of these patients (73%) had polytrauma with at least one more body region injured with abbreviated injury scale (AIS)> 2, besides pelvic trauma. The most common associated injuries were chest injuries (34%) and extremities injuries (30%). During the primary survey, 23 patients were FAST positive, and only 11 of them were found to have associated intraabdominal injuries at damage control laparotomy. However, only 4 out of 11 patients had significant injuries (3 patients with > grade 3 liver injury and one patient with shattered spleen) that can contribute to hemorrhagic shock. Overall mortality was 64%. Twenty-nine patients (43.9 %) died within the first 24 hours of injury; 27 patients due to unresponsive hemorrhagic shock and two patients succumbed to severe head injuries. Seven patients died within ten days of surgery: three due to persistent hemorrhagic shock, two due to ARDS, and two succumbed to MODS. Six patients died due to sepsis-related complications after a hospital stay ranging 10 to 77 days.

Various parameters were compared between the survived and the deceased patients, excluding two patients, who were having severe head injury as primary cause of death (Table 2). Both groups were comparable in terms of mean age and ISS. Mean systolic BP was significantly low in the deceased group, and they received more packed red blood cells (PRBCs) and platelet transfusions than the survived group.

**Table -1.** Demographic profile, mechanism of injury, vital parameters, and outcome data of the study population.

| Parameters | Results |
| --- | --- |
| Total no. of patients | 66 |
| Male, n(%) | 55(83.3%) |
| Age in years, mean±SD | 36.12±12.97 |
| Mechanism of injury, n(%) |  |
| Road traffic injuries | 43(65.2%) |
| Fall from height | 12(18.2%) |
| Train associated injuries | 8(12%) |
| Fall of a heavy object | 2(3%) |
| Others | 1(1.5%) |
| Vital signs at admission |  |
| Systolic BP in mmHg, mean ± SD | 77±34.46 |
| Heart Rate, mean ± SD | 114±19 |
| Class I HS, n(%) | 5(7.6%) |
| Class II and above HS, n (%) | 61(92.4%) |
| GCS at admission, mean ± SD | 12.59±4.07 |
| ISS, median (IQR) | 30 (26-36) |
| Number of patients with ISS ≥ 25, n (%) | 12(18.1%) |
| Disposition time from ED to OR in minutes,<br>Median(IQR) | 30(15-60) |
| FAST examination |  |
| Positive, n(%) | 23(34.8%) |
| Negative, n(%) | 43(65.2%) |
| Blood transfusions in the first 24 hrs. |  |
| PRBCs, median (IQR) | 8.5 (6-12) |
| FFPs, median (IQR) | 4.0 (4-10) |
| Platelets, median (IQR) | 4.0 (4-10) |
| LOS in ICU, in days, median(IQR) | 2.5(1-11) |
| LOS in hospital in days, median(IQR) | 3.5(1-19) |
| Mortality, n(%) | 42(63.6%) |
| Number of patients died within the first 24 hours, n(%) | 29(43.9%) |

**Table-2.**
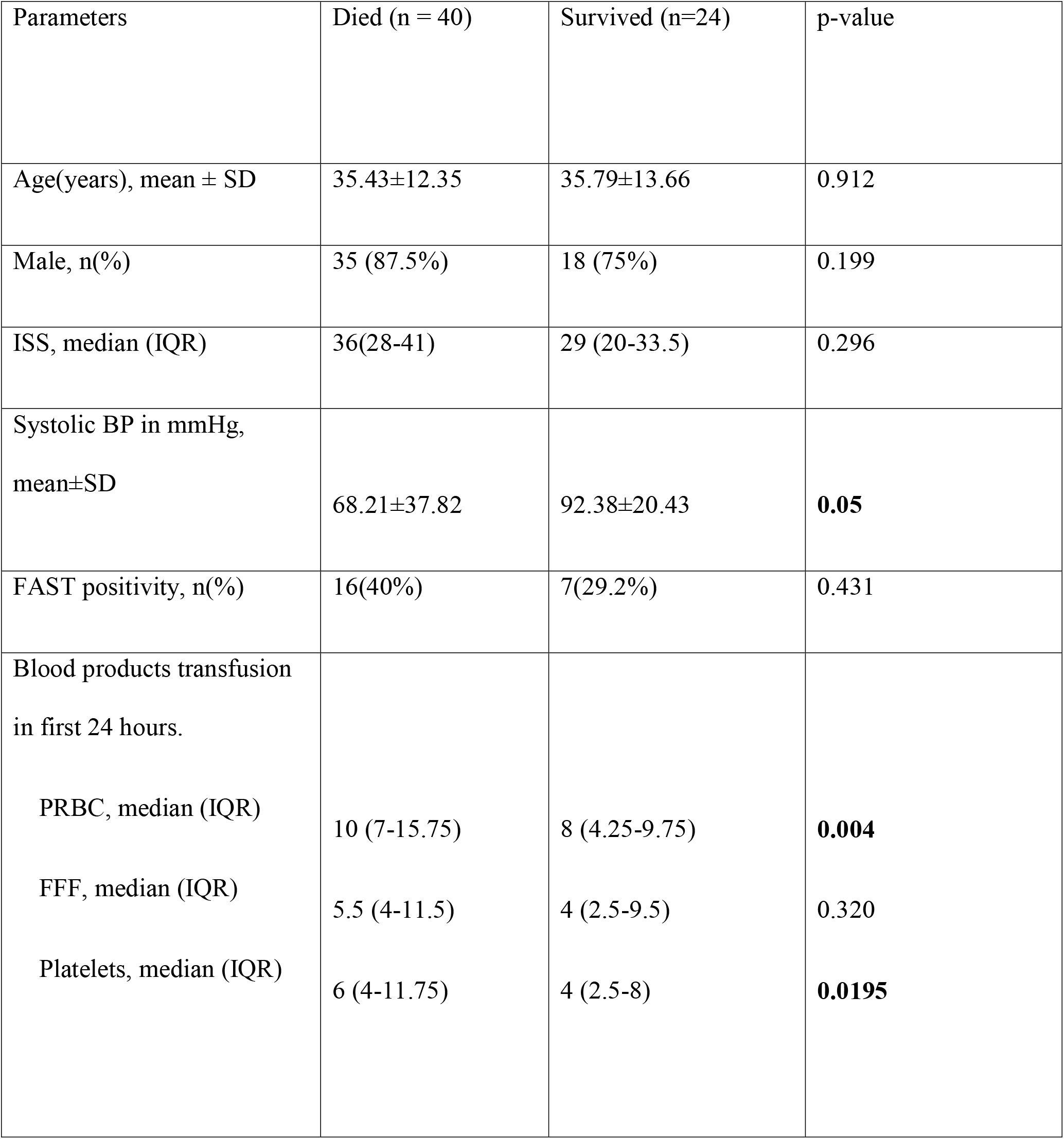
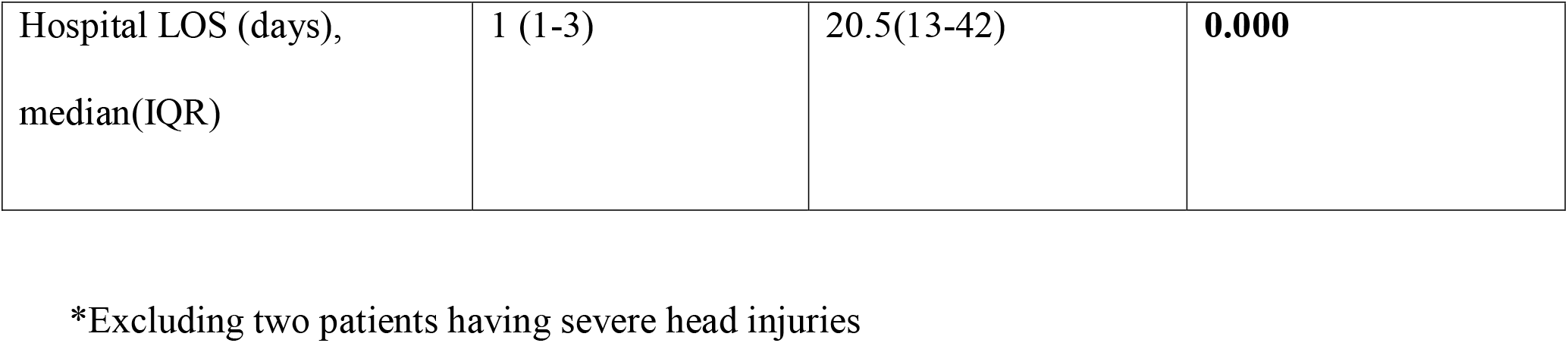
Comparison of parameters between survived group and deceased group*.

| Parameters | Died (n = 40) | Survived (n=24) | p-value |
| --- | --- | --- | --- |
| Age(years), mean ± SD | 35.43±12.35 | 35.79±13.66 | 0.912 |
| Male, n(%) | 35 (87.5%) | 18 (75%) | 0.199 |
| ISS, median (IQR) | 36(28-41) | 29 (20-33.5) | 0.296 |
| Systolic BP in mmHg,<br>mean±SD | 68.21±37.82 | 92.38±20.43 | **0.05** |
| FAST positivity, n(%) | 16(40%) | 7(29.2%) | 0.431 |
| Blood products transfusion<br>in first 24 hours. |  |  |  |
| PRBC, median (IQR) | 10 (7-15.75) | 8 (4.25-9.75) | **0.004** |
| FFF, median (IQR) | 5.5 (4-11.5) | 4 (2.5-9.5) | 0.320 |
| Platelets, median (IQR) | 6 (4-11.75) | 4 (2.5-8) | **0.0195** |
| Hospital LOS (days),<br>median(IQR) | 1 (1-3) | 20.5(13-42) | **0.000** |

\*Excluding two patients having severe head injuries

## Discussion

Pelvic fracture occurs when a significant amount of blunt force is transmitted to the body and, therefore, is often associated with injuries to other vital organs. Pelvic fractures themselves can cause significant morbidity and mortality because of massive hemorrhage. Vital signs at presentation and response to initial resuscitative measures are suggestive of massive blood loss (9). All our patients were non-responders after the primary survey. The amount of blood transfusion can also be used as a surrogate marker for the massive blood loss and hemodynamic instability. A multicentric trial showed that the patients presenting with hemodynamic instability received a median of 7.5 units of PRBC, 6 units of fresh frozen plasma (FFP), and 3.5 units of platelet transfusions (10). Our patients received a median of 8 units of PRBC, 4units of FFP, and 4units of platelet transfusions during the first 24 hours of resuscitation.

The mortality rate due to pelvic fractures in hemodynamically unstable patients ranges from 40% to90% (11). Various treatment modalities for control of hemorrhage such as PCCDs, C-clamp, external fixator, preperitoneal pelvic packing (PPP), angioembolization (AE), bilateral ligation of the internal iliac arteries (BILLA), and resuscitative endovascular balloon occlusion of aorta (REBOA) have been reported (12). Management protocols differ widely across the globe and primarily depend on the availability of resources. A PCCD application either in prehospital setting or in ED is a crucial step in the management of a patient with pelvic fracture. Source of blood loss is venous in around 85% of patients with an unstable pelvic fracture and arterial in the rest 15% patients (13). Proper use of PCCD reduces open-book type fractures, decreases volume of the pelvic girdle, and can also produce a tamponade effect sufficient enough to stop venous bleeding (14). External fixation devices are also used for this purpose, but their application needs a variety of appropriate instruments, intra-operative imaging, and expertise. This may lead to an unacceptable delay in the management of hemodynamically unstable patients (15). Moreover, there is no advantage of external fixator over PCCD for immediate hemorrhage control. For the same reason, we do not use external fixator at the first place in ED. We routinely use bed sheets as PCCDs in all patients with pelvic fracture in ED.

Currently, a two-pronged approach has been suggested for the definitive control of the pelvic fracture bleeding - PPP to control venous bleeding, and angiographic embolization to control arterial bleeding. PPP has stood the test of time in the majority of such patients. Cothren et al. reported better outcomes with early pelvic packing in hemodynamically unstable patients with pelvic injuries in comparison to early AE (16). In another study, Osborn et al. concluded that pelvic packing is fast and as effective as pelvic AE, for achieving hemodynamic stability (17). They reported that median disposition time from ED to OR in PPP group was 45 minutes and 130 minutes in angiography group. Median disposition time from ED to OR in our study was 30 minutes (IQR; 15-60). Despite its advantages, PPP is not routinely employed. Only about 30% of trauma surgeons in the United States of America perceive packing as an effective means, and nobody prioritizes packing over AE (18).

AE has been more frequently used to control pelvic fracture-related bleeding, especially in cases where the mechanical compression by pelvic packing and PCCDs were ineffective (19). Although the literature has shown promising results with AE, but it does not provide clear indications for the same. Therefore, the decision to do angiography should be guided primarily by the hemodynamic status of the patient. Recent literature suggests that early AE, along with PPP, are complementary techniques, and should be used together to achieve definitive haemorrhage control (12,20). Availability of a hybrid OR is very crucial to perform AE and PPP simultaneously. But even in most advanced trauma centres with hybrid facilities, angioembolisation may take 3 to 5 hours to accomplish (21-22).

BLIIA is an established lifesaving procedure to control massive pelvic bleeding in obstetric and gynecological procedures (23). Miller first extrapolated this technique in patients of pelvic fractures with uncontrolled bleeding and recommended its use in such patients (24). Many authors have reported their experience with this technique in patients with pelvic injury. Jun et al. described BLIIA in twenty-one patients for exsanguinating hemorrhage control (8). With the growing experience of managing these patients worldwide, it became clear that control of bleeding in such patients needs combined strategies to control both arterial and venous bleeding. Dubose and colleagues reported their experience of BLIIA and PPP with a mortality of 64% (7). They concluded that BILLA with PPP was a valuable option in patients with exsanguinating bleeding from pelvis injuries. Our study showed comparable results with a mortality of 64%. There is a concern about losing tamponade effect as in our technique, we approaching to preperitoneal space via intraperitoneal route. As the BLIIA is performed along with, the possibility of further expansion of hematoma is mitigated and PPP will create adequate pressure to stop venous bleeding. BLIIA and PPP may be used as an effective alternative to AE in resource constraint settings.

## Conclusion

Pelvic fractures can cause massive blood loss. Multimodality treatment is required to reduce associated mortality. BIIAL with PPP is an effective and viable alternative to AE with PPP for hemorrhage control in resource constraint settings.

## Data Availability

all analysed data are available and may be made available if needed.

## Authorship statement

DB, MA, NB, SK, and NC participated in the study conception, study design, and analysis of data. DB, SK, AK, PP, JA, AG, BM, and SS contributed to the drafting of the article. All authors participated in critical revisions.

